# A study protocol for a site-randomized trial of a multi-level digital intervention to reduce maternal morbidity & mortality

**DOI:** 10.1101/2025.10.29.25339066

**Authors:** Sydney S. Kelpin, Claire E. Margerison, Athena S. McKay, Elizabeth Vickers, Mary K. Crawford, Jessie Spencer, Alla Sikorskii, Steven J. Ondersma

## Abstract

**Introduction:** Compared to other high-income countries, the United States continues to have the highest rates of pregnancy-related and associated mortality and morbidity (PRAMM), particularly in rural areas and among non-Hispanic Black pregnancies. Over 80% of pregnancy-related deaths are preventable; however, the intensity of existing interventions has proven difficult to broadly disseminate. Technology offers the potential to address such barriers. This study will develop a multi-level digital intervention to reduce PRAMM and evaluate its effects using a site-randomized trial.

**Methods and analysis:** The Michigan Healthy Mom (MI MOM) intervention will be developed using a community-partnered approach and will seek to address PRAMM risks at four distinct levels: individual, support system, provider, and community.

Pregnant participants and up to three members of their personal support system will receive an initial brief interactive session through a mobile web app and will thereafter receive a series of text messages with links to extended content. Healthcare providers will receive biweekly text messages and/or flyers distributed in clinic staff areas, and community health workers (CHWs)— who can facilitate access to local services—will be available via secure live chat text access. MI MOM effects will be evaluated using a cluster randomized trial in 10 antenatal care clinics throughout Michigan (*N* = 500 pregnant participants age 18+ receiving Medicaid). We will compare intervention and control arms on two co-primary outcomes: total PRAMM through 1 year postpartum as measured using a universally collected linked dataset of Medicaid claims and vital records and an index of PRAMM risk factors directly targeted by MI MOM.

**Ethics and dissemination:** The Michigan State University (MSU) Institutional Review Board (IRB) has provided ethical approval (STUDY00011005). Results will be disseminated via presentations at academic conferences and community forums, as well as publications in peer-reviewed journals.

**Trial registration number:** ClinicalTrials.gov Registry (NCT07213284).

**STRENGTHS AND LIMITATIONS OF THIS STUDY:**

• Community co-creation of the intervention content ensures MI MOM reflects the best available evidence-based practices while also being deeply rooted in the preferences and needs of the intended users.
• The technology-driven approach provides a unique level of scalability for the developed intervention, enabling MI MOM to be readily implemented in practice with high fidelity and minimal training across contexts.
• Integrating technology-delivered services with CHW live chat text access allows on-demand connection to helping professionals to facilitate warm handoffs to locally available services.
• Digital approaches can elicit concerns around offering a minimally intensive approach to addressing the complexity of PRAMM; the ideal public health response would include person-delivered approaches used in concert with highly scalable, low-barrier technology-driven programs.

## INTRODUCTION

Pregnancy-related and associated mortality and morbidity (PRAMM) is a major public health concern in the United States (US). Compared to other high-income countries, the US continues to have the highest rate of pregnancy-related mortality with 669 maternal deaths reported in 2023 [1,2]. For every pregnancy-related death, approximately another 70 women experience severe maternal morbidity (SMM), defined by the Centers for Disease Control and Prevention as “unexpected outcomes of labor and delivery that can result in significant short- or long-term health consequences” [3]. The annual rate of SMM in the US increased by over 20% between 2008 and 2021 [4]. Moreover, maternal deaths due to homicide, suicide, and drug overdose have been identified as leading causes of pregnancy-associated deaths and have also increased substantially in recent years [5].

Black women are three times more likely to die from pregnancy-related causes [6] and approximately twice as likely to experience SMM compared to their non-Hispanic (NH) white counterparts [7]. Further, Black women have nearly 5 times the risk of pregnancy-associated death due to homicide compared to NH white women [5]. PRAMM outcomes have also been found to vary by region, with those living in rural areas at increased risk of SMM and mortality compared to those living in urban communities [8,9]. Recent research also suggests that certain risks related to age and race may be even more pronounced in rural areas [9]. Finally, rates of substance use during pregnancy have also been found to vary by region, with pregnant women in rural communities reporting higher rates of substance use compared to urban women, as well as higher rates of neonatal abstinence syndrome and other associated adverse birth outcomes [10–13].

Over 80% of pregnancy-related deaths are preventable [14] and represent an opportunity for prevention and intervention efforts. Due to the complex nature of PRAMM, such efforts require an intensive multilevel approach that addresses contributing factors at the community, provider, family/support system, and individual level [15]. However, highly intense interventions and initiatives can be difficult to broadly disseminate due to the required state-level support, funding, expertise, training, and community involvement [16,17]. Further, rural areas and minoritized communities also often see less benefit from such programs due to barriers around distance, service availability, and other key resources [18,19].

Technology offers the potential to address such barriers and provide a vehicle for addressing PRAMM risks. Digital interventions can be readily deployed in clinics at low cost and require less time and training of personnel compared to person-delivered approaches [20,21]. Technology can also address specific patient barriers to accessing and utilizing perinatal care, including provider distance and availability, difficulty with appointment scheduling, transportation, childcare, and time off work [22–25]. Further, due to the high rates of smartphone ownership among young adults (98% of those age 18-29) [26], technology offers broad reach to patients that remains readily accessible wherever they go. Finally, recent research also suggests technology has the potential for reducing PRAMM by improving adherence to clinical guidelines, improving access in rural areas, and facilitating identification of risk [27].

The proposed study was designed in collaboration with community partners to test the efficacy of a multi-level, text-messaging focused digital intervention to reduce maternal morbidity and mortality. The intervention will address key PRAMM risks using a multilevel approach, with further optimization to address the specific needs of Black pregnant women and those residing in rural areas. In addition, it will provide pregnant participants with immediate live chat text access to a Community Health Worker (CHW) who can facilitate connection to local services. Specific aims are to: 1) collaborate with pregnant and postpartum women, healthcare providers, family members, and researchers—particularly those identifying as Black and those from rural areas—to develop the Michigan Healthy Mom (MI MOM) app, with specific content for pregnant participants, their personal support system and providers; and 2) evaluate MI MOM effects on PRAMM using a cluster randomized trial.

## METHODS AND ANALYSIS

### Study design

This study is a cluster randomized trial involving 10 clinics throughout Michigan randomized to receive treatment as usual (TAU), which includes an electronic single-session screening and brief intervention app, or TAU plus the MI MOM app and text messages with CHW live chat access. This study was designed as one of three projects that make up the Maternal Health Multilevel Interventions for Racial Equity (MIRACLE) Center (U54 HD113291), one of the Maternal Health Research Centers of Excellence funded through the National Institutes of Health’s (NIH) IMPROVE Initiative [28]. This study was designed to rigorously test the efficacy of a digital intervention to reduce PRAMM while accounting for the effects of interventions administered as part of other projects in the Center at the county level. For this reason and to prevent contamination across participants seen in the same clinic, cluster randomization will occur at the level of the county, with one clinic per county. The first co-primary outcome is a total PRAMM index (count of mortality and morbidity conditions) through one year postpartum as measured using a linked dataset of Medicaid claims and vital records.

The second co-primary outcome is a self-reported index of PRAMM risk factors directly targeted by MI MOM. It will be measured repeatedly at baseline, and then 5 more times: 6 weeks post-baseline and 35 weeks gestation, and the first postpartum year (6 weeks, 6 months, and 12 months postpartum). The primary analysis will focus on evaluation of the main intervention effect on the two co-primary outcomes. We will evaluate if the intervention effects on the Medicaid claims-based PRAMM index are mediated by the proximal outcome of self-reported index of PRAMM risk factors directly targeted by MI MOM. The secondary analysis will focus on differential effects of the intervention according to rural versus urban residence and other potential moderators. This study is a pragmatic trial with non-restrictive eligibility criteria, flexibility in intervention delivery, use of a real-world setting, and a focus on clinically relevant outcomes. The study is expected to begin in November of 2025 and to end in June of 2029. See Figure 1 for a trial overview.

**Figure 1.**
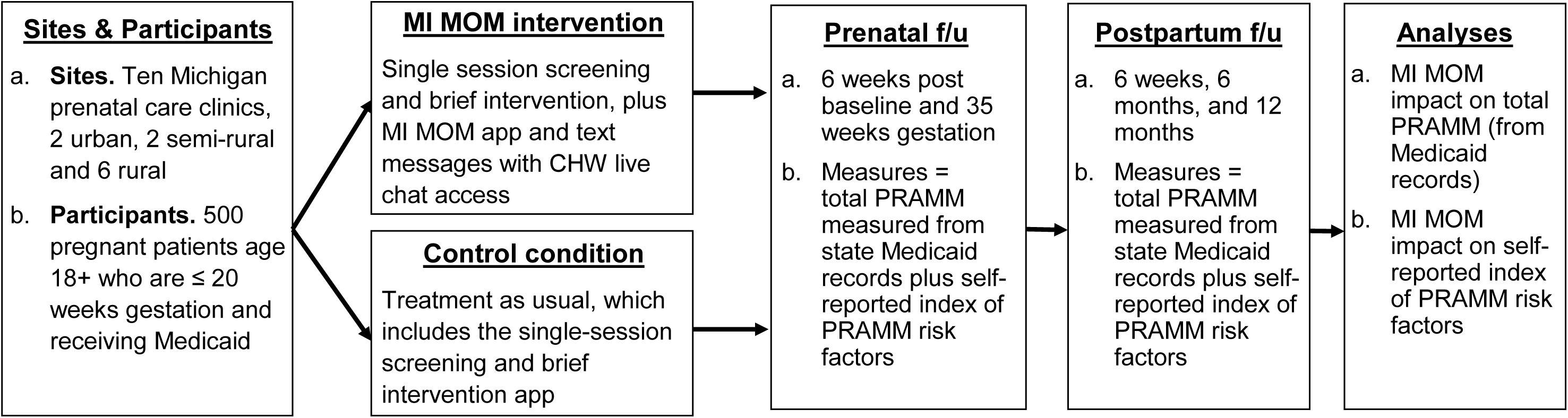
MI MOM trial overview.

### Patient and public involvement

The MI MOM app and text messaging content will be developed in close collaboration with community partners via several different methods throughout the project. First, a Community Advisory Board (CAB) consisting of representatives of the Flint community and rural areas in Northern Michigan will meet regularly throughout the app development phase to provide guidance on content, resources, and overall design and layout. Two collaborative content development events (“Appathons”) will also be held at which content will be developed directly by community members. The MI MOM app will be built using the flexible, no-code Computerized Intervention Authoring System (CIAS) 3.0 platform [29], which enables community members to directly create intervention content that will be incorporated into the final version of app. Finally, the MI MOM app content will be iteratively modified and refined in response to ongoing feedback from the CAB, pregnant patients, family/support system members, and providers prior to trial initiation.

### Setting and participants

Clinics will be drawn from an existing network of antenatal clinics throughout Michigan already using an electronic screening and brief intervention app at the new pregnancy intake appointment [30]. We expect that two clinics will be from urban counties, two from semi-rural counties, and six from rural counties of Michigan (total N=10 clinics). Rural counties will be defined according to census data and the Federal Office of Rural Health Policy guidelines, including non-metropolitan counties and outlying metropolitan counties with no population from an urban area of 50,000 or more people [31]. Semi-rural counties will be defined as Metropolitan counties that include sub-county units that have been designated as rural [32].

Participants in the trial will include pregnant women and members of their personal support system. We will recruit ∼50 pregnant participants from each clinic (total *N* of 500). Inclusion criteria include being pregnant, ≥18 years of age, <20 weeks gestation, and Medicaid covered/eligible. Exclusion criteria include inability to communicate in English and not owning a smartphone with text messaging and data. In addition, we will also include up to three adult members of the index participant’s social support system, to include family, friends, and/or partners. Social support participants will also be required to speak English and own a smartphone. Finally, healthcare providers at each of the ten participating clinics (obstetricians as well as family practice physicians and nurse midwives; n∼30 total) will also be included if they agree to receive text messages and view the brief content provided through those text messages. For all intervention condition sites, we will provide PRAMM-related flyers and posters in the staff areas of clinics and will refresh those flyers and posters at regular intervals.

### Sample size determination

Given the sample size of 10 clinics with an average of 50 pregnant participants per clinic, and an intraclass correlation of .003 estimated based on 2019 Michigan Department of Health and Human Services (MDHHS) Data Warehouse data, we used design-effect adjusted N=436 to determine power in two-sided tests. Because two co-primary outcomes were specified in advance, .05 level of significance will be used for the analyses of both, with the Medicaid claims based PRAMM index getting the higher strength of interpretation. For this outcome there will be almost no missing data, and power is sufficient (.80) to detect the effect size of Cohen’s d=0.27 for between-arm differences. For the outcome of self-reported composite intervention target index, 5 repeated measures will be analyzed with covariance adjustment for baseline version of the outcome. We accounted for 20% attrition and used an N=348 attrition and design-adjusted sample size for conservative power considerations based on classical analysis of repeated measures. We assumed correlation coefficient r=.4 between pairs of repeated measures of the index to estimate reduction in variance and power. Based on the preliminary data, we expect differences between trial arms of Cohen’s d=0.35. With the planned sample size, power exceeds .90 for effect sizes greater than d=0.25 for this outcome. Because of control for clinic location in randomization, the cells represented by the means will be nearly equal in size, and using SAS PROC GLMPOWER, the detectable effect size is d=0.37 for the target intervention index and d=0.54 for PRAMM based on Medicaid claims data. The effect size of half of the standard deviation is a well-accepted threshold for clinical significance [33,34], making clinically significant differences in improvement in primary and secondary outcomes detectable with the planned sample size.

### Randomization

Randomization will occur at the county/clinic level prior to participant recruitment. Sites will be randomly assigned to receive services as usual or the MI MOM app and text messages with CHW live chat access. Clinics will be blocked into blocks of size 2 to achieve the same distribution of rural/urban counties as well as factors related to additional studies under the MIRACLE Center unrelated to the present intervention. Allocation concealment will be facilitated by the randomization being performed by a statistician unaffiliated with study sites. Medicaid claims-based PRAMM-related outcome will be extracted from a universally collected statewide linked dataset, and the staff who maintain this dataset will not be informed of study conditions. All self-reported outcomes will be collected via direct participant report using online survey software. In addition, all data analysts will be blinded to study conditions.

### Recruitment

Pregnant participants will be recruited through an existing electronic screening and brief intervention app [30], which is completed prior to new obstetric intake appointments at prenatal care clinics across Michigan; or through flyers at Michigan prenatal care clinics. Participants indicating interest and meeting eligibility criteria will view a brief description of the study and be asked if they are interested in hearing more. Study staff will contact those who agree by phone to describe the study in more detail and answer questions. Those who agree to participate will be sent a link to the electronic consent and baseline survey and—for those from intervention condition sites—to the initial MI MOM session. After providing consent, participants from intervention condition clinics will be asked to provide the names and cell phone numbers of up to three key support people in their lives (e.g., partner, family member, or friend), with the understanding that they will be sent a link inviting them to participate in the study. Those support persons who click the link indicating potential willingness will be taken through a brief online description of the project and shown a full online consent form; those who provide consent will begin receiving text messages.

Healthcare providers from intervention condition clinics will be asked to give consent, also using an online CIAS 3.0 link, prior to site randomization. As this is a pragmatic trial, provider consent will not affect clinic inclusion or randomization. Provider participants from intervention condition clinics who agreed to be part of the study will begin receiving text messages following randomization. For all clinics allocated to the intervention condition, study staff will provide a series of flyers and posters with PRAMM-related content, which will be renewed with new materials quarterly.

### Intervention

The MI MOM app will operate at four distinct levels, with multiple specific techniques at each level (see Figure 2). These intervention strategies will take place in addition to the current single-session app addressing substance use and mental health. Content will be delivered through weekly text messages sent throughout pregnancy and the first year postpartum, offering proactive content to participants rather than requiring them to seek it out.

**Figure 2.**
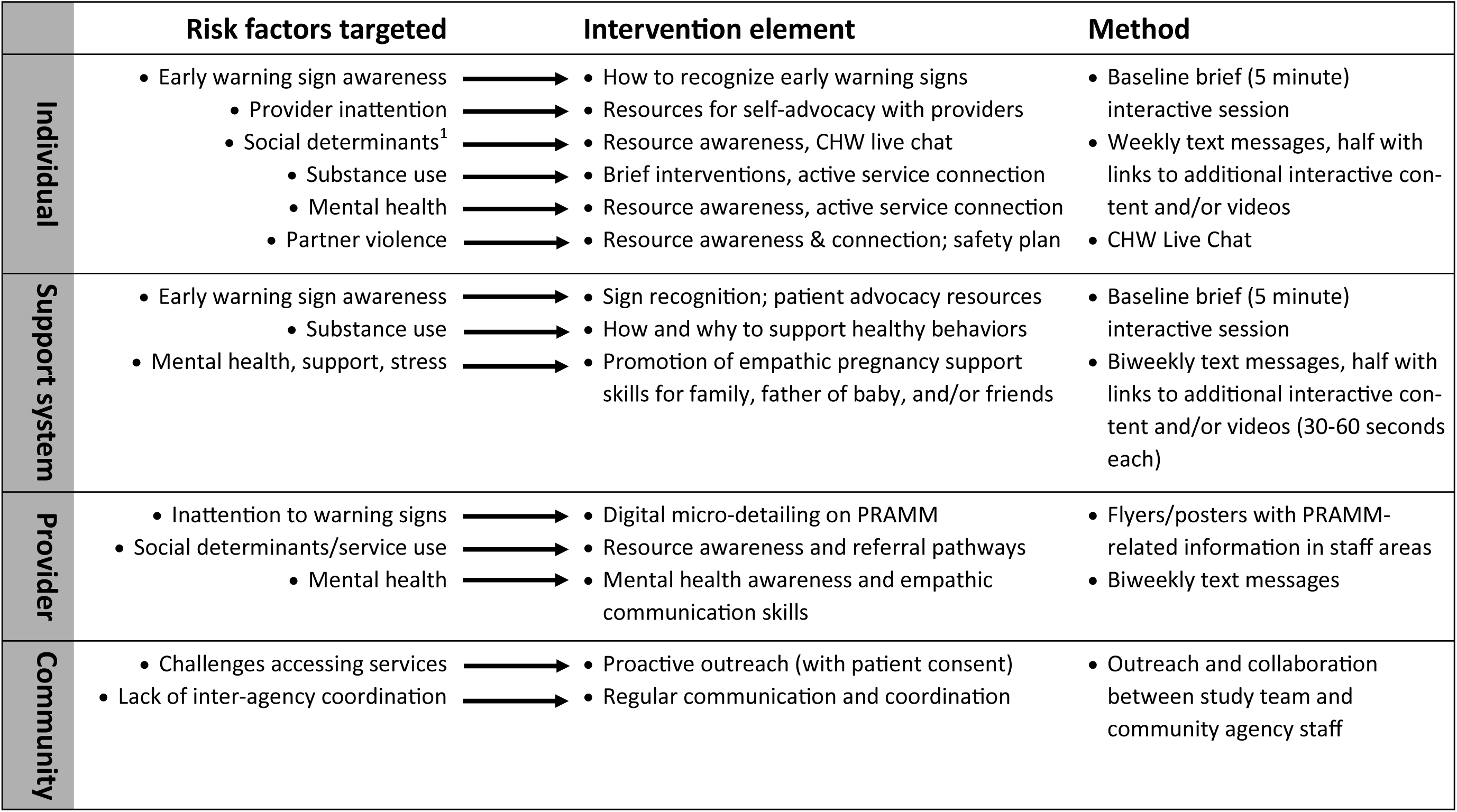
MI MOM intervention overview. ^1^ Housing, transportation, child care, access to care, etc.

#### Community level

Community-level efforts will focus pragmatically on promoting use of Medicaid’s evidence-based [35–37], home visiting program (MIHP), which in Michigan is available in all regions, but is at present only used by 20% of eligible pregnant women. Efforts at this level will also focus on facilitating “warm handoffs” directly from MI MOM to local agencies and treatment providers, especially through the live chat feature. These “warm handoffs” will involve either seeking participant permission to have someone from the home visiting or other agency proactively reach out to the participant, and/or live three-way phone calls to initiate services. We will work closely with major programs in each area seeking arrangements to facilitate direct connections with services.

#### Provider level

All participating clinics that are allocated to the intervention condition will receive flyers and/or posters that will be displayed within staff areas. These materials will provide PRAMM- related information, particularly regarding patient mental health and social determinants of health. These materials will be refreshed quarterly. Providers at participating sites will also have the option to receive one text message with PRAMM-related content every two weeks. Content for providers will include prompts to provide support for early home visitation and other service involvement, reminders of key risk factors for PRAMM, and education on empathic patient communication.

#### Support system level

After providing consent, support persons will begin receiving text messages every two weeks. As with the pregnant participants, these messages may be text only or may include a link to extended content such as a brief video or a brief interactive intervention. Content will focus on areas such as how to be empathic and supportive, how to help recognize warning signs, and how to support healthy behaviors.

#### Individual level

Pregnant participants will complete an initial interactive session that will describe the rationale for MI MOM and the process of weekly incoming text messages. Weekly texts thereafter will emphasize key PRAMM reduction messages on topics listed in Figure 2. These messages may be text only, text with a link to specific resources, or a text with links extended content such as a brief video or interactive app session (60-90 seconds). The MI MOM app will also provide tools for pregnant and postpartum women to advocate for themselves regarding their concerns and symptoms. While interacting with the app, pregnant participants will see an icon in the lower right corner that they can click for an instant and secure text chat connection with a CHW to facilitate connection to locally available services. The availability of this service will also be emphasized, along with a link to the live chat service, throughout the text message cycle. See Figure 3 for illustration of an example text message and interactive app session.

**Figure 3.**
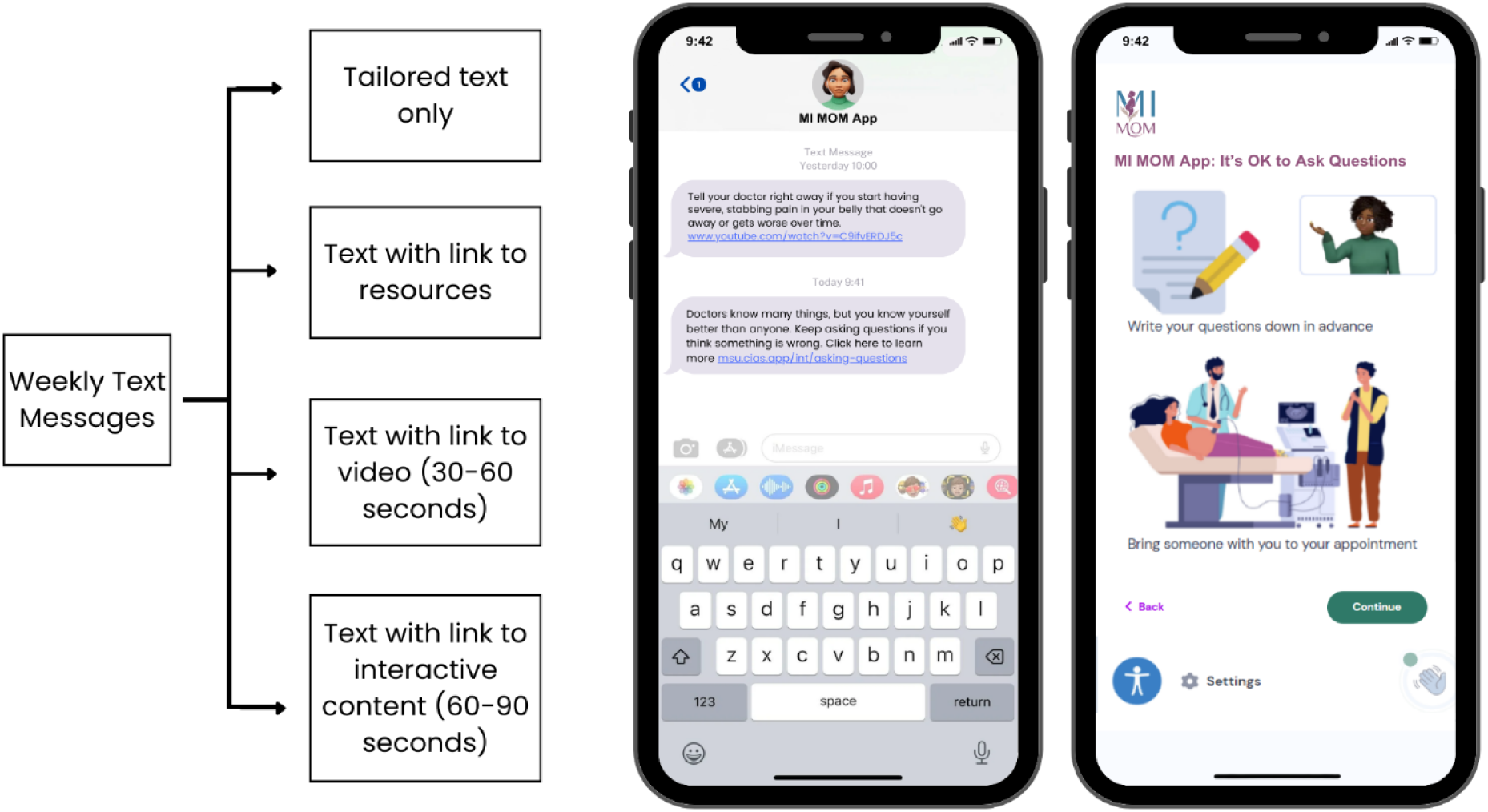
Illustration of MI MOM for pregnant participants.

### Control Condition

Sites randomized to the control condition will receive usual care, including the electronic screening and brief intervention app that is already integrated with antenatal services throughout Michigan. This comparison group was chosen to examine the extent to which adding the MI MOM app and text messages can improve upon treatment as usual.

### Procedures

#### Text messaging

After providing consent, participants (pregnant, social support persons, and providers) will complete a 5-minute brief interactive session introducing them to the intervention and a rationale for the focus on maternal morbidity and mortality. This introduction will include the content developed during the “Appathon” community events, with urban sites seeing content developed during the urban Appathon and rural sites seeing content developed during the rural Appathon. After completion of this introductory session, participants will begin receiving weekly text messages, which will be the core means by which they receive MI MOM intervention content, as well as requests for follow-up data.

Each intervention text message will be sent once. A “STOP” request from a participant will cease messages, and result in a phone call to ensure that the participant knows how to restart text messages, if they choose to. Intervention participants who choose to end text messages will continue to receive messages for follow-up data collection. Text messages with links to follow-up data collection will be sent up to three times each, followed by a single phone call if needed. CHW and live chat

Participants who click for live chat (texting) with the CHW will be immediately connected if a CHW is logged in. If a CHW is not logged in, the participant will receive a message saying that the CHW has been notified that someone is requesting support and provided resources (e.g., 988 Lifeline). The CHW will use detailed information regarding local resources to facilitate connection to services through one of three methods: (a) having the CHW call the program of interest to arrange an appointment time during the chat; (b) sending the service agency the participant’s consent and contact information, allowing the agency to reach out; or (c) obtaining participant permission for the CHW to call the referral program and initiate a 3-way phone call with the participant once they are connected. The CHW will also provide short-term support as needed, following a manual with clear guidance in responding to suicidality or other immediate risks. The CHW will be trained in adherence to the manual, with regular supervision and review of chat transcripts.

#### Participant Compensation

Pregnant participants will be compensated for each assessment completed at baseline, during pregnancy (3 months post-baseline and 35 weeks gestation) and during the first year postpartum (6 weeks, 6 months, and 12 months). Participants will receive a $50 electronic gift card for all follow-up sessions, other than the 12-month postpartum follow-up, for which they will receive a $100 gift card. Social support participants will receive $50 for completing initial study screening questions when they enter the study, $10 at 35 weeks gestation, and $50 one year after the baby is born. Participants will only be compensated for data collection and not for intervention activities. Provider participants will not receive any compensation for their participation.

### Measures

Data on the first co-primary outcome will be extracted from the MDHHS Data Warehouse. Linked sources of data, all routinely collected or publicly available (e.g. census tract data), include: (1) complete Medicaid pregnancy and postpartum (12 months after birth) medical claims (2) monthly Medicaid eligibility during pregnancy and postpartum; (3) birth records, including data on pre-pregnancy and pregnancy risk factors (e.g. chronic disease, prior preterm births); (4) maternal death records; (5) census tract data; and (6) some prenatal and postpartum screening data.

The second co-primary outcome of participant report on proximal intervention targets of the MI MOM app (see Figure 2) will be measured repeatedly throughout pregnancy (6 weeks post-baseline and 35 weeks gestation) and the first postpartum year (6 weeks, 6 months, and 12 months postpartum). Standardized measures will be used for risk categories when available, including the Tobacco, Alcohol, Prescription medication, and other Substance use Tool for substance use (TAPS) [38,39]; the Humiliation, Afraid, Rape, Kick questionnaire for intimate partner violence (HARK) [40], and the Patient Health Questionnaire (PHQ-9) [41] and Generalized Anxiety Disorder Screener (GAD-7) [42] for symptoms of depression and anxiety. Other domains covered in the index include questions to assess patient understanding of early warning signs and appropriate responses (e.g., calling doctor, emergency room), overall confidence in patient provider communication, and social determinants of health.

Secondary outcome measures will be collected on key maternal health constructs, as well as satisfaction and implementation of the MI MOM intervention. Measures will include assessment of infant care and feeding practices using three items from the Centers for Disease Control Pregnancy Risk Assessment Monitoring System (PRAMS) [43], as well as information on financial strain, food security, and access to medical care. Participant satisfaction with the MI MOM intervention will be assessed using a 6-item scale where participants rate the extent to which they liked the MI MOM app, found it easy to use, respectful, bothersome, and helpful on a 1-5 scale (1 = *not at all* and 5 = *very much*). Lastly, implementation of the MI MOM app will be measured using the 4-item Acceptability of the Intervention Measure (AIM) and the 4-item Intervention Appropriateness Measure (IAM) [44]. See Table 1 for a schedule of measures and assessment time points.

**Table 1.**
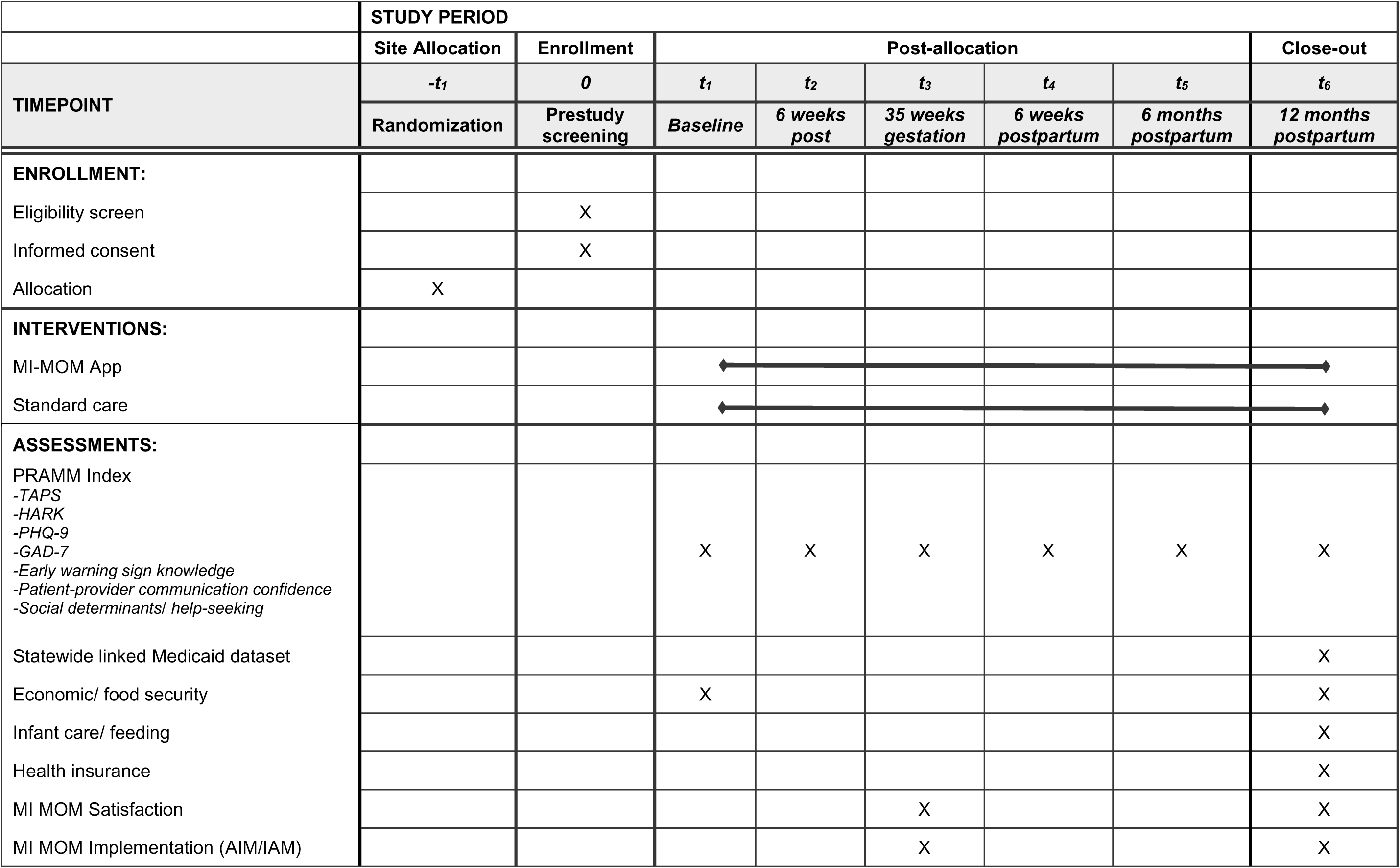
Schedule of enrollment, interventions and assessments.

### Data Analysis

The study will adhere to the Consolidated Standards of Reporting Trials statement for cluster randomized trials [45]. Distributions of outcomes will be summarized, and outliers will be investigated to examine their influence on the results. All analyses will follow the intent-to-treat approach. Missingness will be examined by trial arm for quantity, type and pattern. If patterns of missing data indicate potential not missing at random (NMAR) mechanisms, then models describing missing mechanisms (e.g., pattern-mixture models) and sensitivity analyses will be employed. For the PRAMM outcome, as of 2023, all postpartum women in Michigan can retain Medicaid insurance for 12 months after giving birth.

#### Primary analyses

Each of two primary outcomes, PRAMM and intervention target index, will be analyzed separately. LME or GLME models will relate the outcome measure *y* to the group assignment variable *x*_1_, outcome at baseline *x*_2_(target intervention index only), race/ethnicity, and rural or urban residence, with clinics included as a random effect. In analyses of the target intervention index, an additional random effect will reflect repeated measures of an individual participant over time. The essential parameter is the coefficient for the trial arm variable (main effect, time averaged for the intervention target index), and test for its difference from zero will be a formal test of intervention effect. In addition to statistical significance, we will gauge clinical significance based on estimated adjusted effect size defined as difference between least square means according to the trial arm divided by the square root of residual variance.

#### Secondary analyses

To test intervention effects related to region, trial arm rural/urban residence interactions (one at a time) will be added to LME or GLME models from the primary analyses. The essential parameter in these models will be associated with the coefficient for the interaction term reflecting differences in intervention effects for rural versus urban residents. Sub-scores of the intervention target index will also be analyzed by specific topic areas (e.g., substance use, mental health). Finally, mediation will be tested using the Preacher and Hayes’ [46] approach to estimate direct and indirect (through the mediator of intervention target index) effects of the trial arm on PRAMM.

### Ethics and Dissemination

The Michigan State University (MSU) Institutional Review Board (IRB) has provided ethical approval (STUDY00011005). This study will adhere to the National Institutes of Health (NIH) Data Sharing Policy and Policy on the Dissemination of NIH-Funded Clinical Trial Information. Accordingly, the deidentified study dataset will be uploaded and stored indefinitely in the NIH data archive. Although we can share data generated by this project, we cannot share the MDHHS Warehouse data accessed by this project, which requires a data use agreement.

Additionally, this trial is registered with the Clinical Trials Registry (NCT07213284) and study results will be submitted to ClinicalTrials.gov in compliance with the Final Rule for Clinical Trials Registration and Results Information Submission. The dissemination plan also includes presentation of results at academic conferences, community events and forums, and publication in peer-reviewed journals.

Informed consent will be obtained from all participants prior to entering the trial. The informed consent will detail all potential risks of study participation, including limited data security of text messages due to the potential for messages to be seen by others who have access to their device and that the messages are not encrypted. Participants will also be informed that the MI MOM app addresses sensitive topics, and while every effort has been made to present topics in a way that does not cause discomfort, they are able to skip any content that makes them uncomfortable.

The study team will follow procedures to maintain participant confidentiality and data security throughout all phases of the research. All self-report data will be backed up on a university secure folder that is HIPAA compliant, and a linking table will be stored in a separate location that is only accessible to certain study staff. Any communication among study team members will use study IDs and no identifiable information. In addition, the MDHHS dataset will be linked to individual study participants and subsequently de-identified using the honest broker method.

A Data Safety and Monitoring Board (DSMB) will be responsible for monitoring this trial. The DSMB will be comprised of at least three individuals independent of the research team who do not have competing interests with the study. The DSMB report will be provided annually to the National Institute of Child Health and Human Development (NICHD) and will include confirmation of adherence to the data safety and monitoring plan, any regulatory or quality issues since the last reporting period, a summary of any adverse events (AEs) that occurred over the past year, any resulting actions or changes to the study protocol, and all new and continuing IRB approvals. Using the FDA definition of serious AES, any AEs that are deemed to be serious and considered at least possibly related to study participation will be reported to the MSU IRB as well as the NICHD project officer within 48 hours.

### Author contributions

Drs. Ondersma, Margerison, and Athena McKay, all Principal Investigators (PIs) on the project, co-led the design of this study and writing of the NIH grant application. Alla Sikorskii led the development and writing of the data analysis plan. All authors contributed to the protocol through writing and/or editing.

### Trial sponsor

This trial was sponsored by NIH/NICHD. The study sponsor did not have any role in the study design, collection, management, analysis, interpretation of data; writing the protocol report; or the decision to submit the protocol report for publication. The study sponsor also does not have ultimate authority over any of these activities.

### Patient consent

Patient consent not required for publication.

### Ethical approval

The Michigan State University (MSU) Institutional Review Board (IRB) has provided ethical approval (STUDY00011005). This trial is also registered with the Clinical Trials Registry, NCT07213284, dated October 8, 2025.

## Data Availability

The manuscript is a study protocol and data collection is expected to begin in November of 2025. Following the study, the deidentified study dataset will be uploaded and stored indefinitely in the NIH data archive

